# Current Gaps in Delirium Recognition and Management: A Cross-Sectional Survey of ICU Physician and Nurse Leaders

**DOI:** 10.64898/2026.02.23.26346839

**Authors:** Michelle Armenta Salas, Annie Zhang, Timothy D. Girard, John W. Devlin, Juliana Barr

## Abstract

**BACKGROUND:** Delirium is common in critically ill adults but often goes unrecognized and undertreated. Little is known about the perceptions of ICU nurse and physician leaders regarding ICU delirium detection and management and the potential role of objective continuous delirium monitoring to facilitate ICU delirium care.

**RESEARCH QUESTION:** What are the perceptions of ICU leaders regarding the current challenges associated with delirium recognition and management and the potential benefits of continuous delirium monitoring?

**STUDY DESIGN AND METHODS:** We conducted a blinded, cross-sectional, electronic survey of ICU leaders across the U.S., including physician directors and nursing managers with ≥3 years of ICU leadership experience. We asked about perceptions of the effectiveness of current delirium clinical assessment tools, current delirium detection and management challenges, and how an objective, continuous delirium monitoring system might impact clinician practice and patient outcomes in their ICU.

**RESULTS:** Among the 81 respondents (62 physicians, 19 nurses), most (76%) reported that recommended delirium assessment tools (CAM-ICU, ICDSC) are used in their ICUs, though there were mixed perceptions on how reliably they are conducted. A majority (63-90%) perceived that current bedside assessments delay and limit the recognition of ICU delirium. Nearly all (89%) agreed an objective delirium monitoring tool would be more clinically valuable than current delirium assessment tools and that it would support real-time, delirium management by clinicians.

**CONCLUSIONS:** ICU leaders perceive that there are limitations to using clinical delirium assessment tools in ICU patients to effectively detect and manage ICU delirium. Most felt that an objective delirium monitor could facilitate delirium detection and potentially expedite appropriate delirium management in patients.

## Introduction

Delirium occurs in up to 80% of critically ill patients and is associated with poor clinical outcomes, including an increased duration of mechanical ventilation and intensive care unit (ICU) length of stay, higher mortality, and reduced long-term cognitive function^1-5^. Current guidelines recommend that all ICU patients be managed with the ABCDEF bundle^6^, which includes performing regular delirium assessments with a validated instruments like the Confusion Assessment Method for the ICU (CAM-ICU)^7,8^ or the Intensive Care Delirium Screening Checklist (ICDSC)^9^, and several non-pharmacologic strategies to reduce delirium (e.g., minimization of deliriogenic medications, protocolized wakefulness and liberation from mechanical ventilation, sleep promotion and early mobilization). Despite evidence that the ABCDEF bundle reduces delirium and improves patient outcomes, bundle implementation and compliance remain sporadic^10-12^.

While ICU nurses acknowledge delirium to be a significant problem, they underestimate its prevalence^13,14^. Nurses also report ICU patients are not routinely assessed for delirium with either the CAM-ICU or ICDSC and that adoption of the ABCDEF bundle in their ICUs is low^10-13^. High workload burden, inadequate training, poor physician backing, and lack of leadership support are often cited as barriers to performing routine delirium assessments and ABCDEF bundle adoption^11-13,15,16^. Objective continuous delirium monitors^17-19^ might address some of these barriers but are not currently widely available.

While ICU nurse and physician leaders have important roles in reducing barriers to delirium detection and management, the perceptions and practices of ICU leaders about delirium detection are not well understood nor are their opinions about the potential roles and benefits of continuous delirium monitoring remain unclear. Given the important role that ICU leaders play in evaluating new technology before it is implemented in the ICU, understanding the perceptions of ICU leaders regarding current delirium management challenges and the value of objective delirium assessment technologies could inform the development and implementation of strategies focused on improving ICU delirium detection. We therefore conducted a cross-sectional survey of U.S. ICU physician and nurse leaders to evaluate their perceptions about delirium recognition and management practices, current unmet needs regarding ICU delirium recognition and management, and objective delirium monitoring technologies.

## Methods

### Study Design and Participants

Survey participants were recruited by a survey-distribution agency (Reckner Healthcare Market Research, Chalfont, PA). The agency handled recruitment, and participants were blinded to the sponsor (Ceribell, Inc., Sunnyvale, CA) and researchers’ affiliations. The survey sought to recruit 80 ICU leaders who met the following eligibility criteria: 1) worked in an adult ICU in the US; 2) served as medical director, department head, nursing director or nursing manager; and 3) had ≥3 years of cumulative experience in any of these ICU decision-making roles. The survey sought to recruit 80% physicians and 20% nurses of whom 20%-30% worked at academic medical centers (AMC) or university hospitals. We excluded participants who did not complete all sections of the survey. Participants were compensated for their participation. The agency only tracked participants who clicked the anonymized invitation link and disabled the link after the target participation quota was met.

### Questionnaire Development

The 23 survey items were initially developed as a market research survey. The subject matter and general topics of the survey were discussed between ICU delirium experts and members from an industry sponsor. The surveyed key areas of interest were: 1) current delirium perceptions and recognition; 2) ICU delirium assessment practices and the perceived effectiveness of current bedside assessment tools; 3) ICU delirium management practices and unmet needs regarding ICU delirium recognition and management, including the benefit(s) of early delirium reduction interventions; and 4) their opinions regarding objective delirium monitoring technologies. For the objective delirium assessment section, participants were provided with two brief videos of two different brandless delirium monitoring tool prototypes: 1) Option 1 continuously monitored the patient and provided a binary alert of delirium presence and 2) Option 2 continuously monitored the patient and provided a continuous measure of delirium occurrence and burden (**Videos 1 and 2**, respectively). Respondents were then asked questions about the perceived usefulness of each monitor. Key demographic parameters were initially collected to ensure each respondent met all survey eligibility criteria. After the initial questionnaire development, we did not conduct additional pilot testing given the survey was sent to clinicians as a part of a market research initiative. The full survey is available in the **e-Appendix 1**.

### Questionnaire Administration

The electronic survey was built and distributed on the Forsta survey platform (PG Forsta, Stamford, CT) by the agency using a double-blind design. While survey participation was optional and all responses were anonymized by the agency, this survey project was not approved by an Institutional Review Board before distribution because it started out as a market research survey.

### Statistical Analysis

We used descriptive statistics to present the demographic characteristics of the respondents and responses to multiple-choice questions, reporting numbers and percentages for categorical variables and medians and first and third quartiles for continuous variables. For Likert scale statements, we summarized the level of agreement and disagreement for key questions, defining agreement as a score of 3 (“Agree”) or 4 (“Strongly Agree”) and disagreement as a score of 2 (“Disagree”) or 1 (“Strongly Disagree”). For questions asking to rank features on a scale of 1 through 10, we grouped scores 8-10 as “highly valuable” for analysis. For items with scores 1-7 used to rank priorities, we considered scores 1-2 a “high priority” and “6-7” a “least priority”. We performed Wilcoxon rank-sum test and Kruskal-Wallis test for continuous variables and the Fisher’s exact test for categorical variables to analyze associations between demographic variables—physician vs. nursing roles, AMCs vs teaching, and non-teaching community hospitals—and responses. All statistical analyses were performed using RStudio Integrated Environment for R software (2025, Posit Software, PBC, Boston, MA). *P ≤*0.05 was considered statistically significant and was adjusted for multiple comparisons if multiple demographic categories were present.

## Results

### Study Respondents

The electronic survey link was enabled from March 17 through April 21, 2025. Study flow and participant dropout rates are shown in **Figure 1**. A total of 388 clinicians accessed the survey invitation link, and 196 passed the initial screening questions. Of these, 56 self-terminated during the rest of the demographic questions, and an additional 57 participants (all identified as ICU medical directors) were terminated due to their quota in the survey already being met. Of the remaining 83 respondents, 81 respondents completed the survey in full (**Table 1**). Of these 81, 77% (N = 62) identified as physicians and 75% (N = 61) worked in community hospitals.

**Table 1.** Respondent’ demographics.

|  |  | N (%) |
| --- | --- | --- |
| Profession | Physician | 62 (77%) |
|  | RN / NP | 19 (23%) |
| Medical Specialty (Physician) | Pulmonary | 13 (21%) |
|  | Critical Care Medicine | 32 (52%) |
|  | Neuro Critical Care | 9 (15%) |
|  | Hospitalist | 8 (13%) |
|  | Others | 0 |
| Type of ICU (multiple selection allowed) | Medical | 70 (86%) |
|  | Surgical | 46 (57%) |
|  | Cardiac | 32 (40%) |
|  | Neuro | 34 (42%) |
|  | Other | 2 (2%) |
| Role in department | Medical Director | 44 (54%) |
|  | Department head | 18 (23%) |
|  | Nursing Director / Manager | 19 (23%) |
| Type of Hospital | Academic Medical Center / University Hospital | 19 (23%) |
|  | Community Hospital, Teaching | 27 (33%) |
|  | Community Hospital, non-teaching | 34 (42%) |
|  | Government Hospital (VA / Military) | 1 (1%) |
| Location of Hospital | Urban | 32 (40%) |
|  | Suburban | 44 (54%) |
|  | Rural | 5 (6%) |

**Figure 1.**
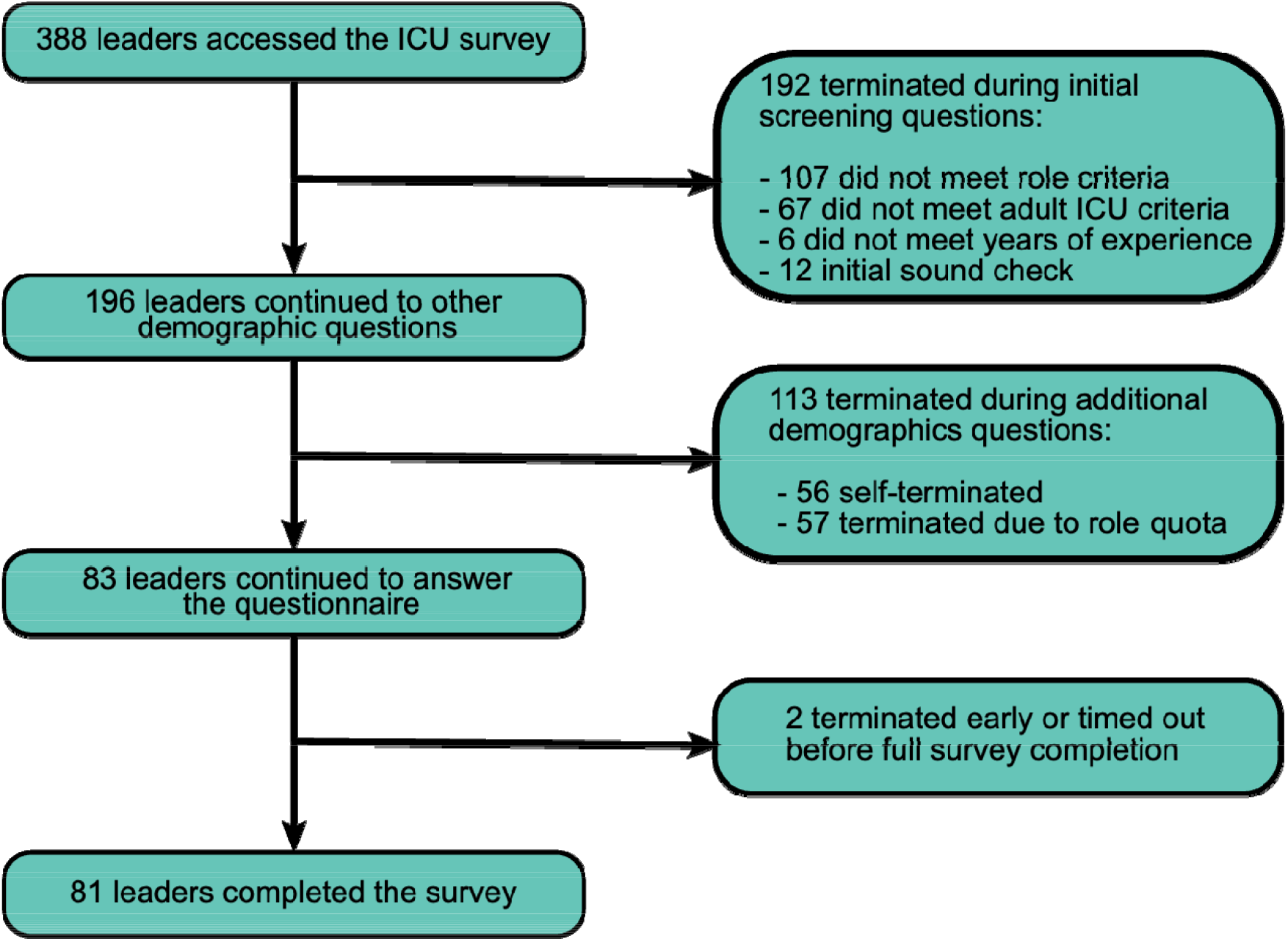
Participants response flow with dropouts/terminations reasons.

### Delirium Prevalence and Detection

Nearly all (94%) respondents agreed that delirium is prevalent in their patients (**Table 2**), although a third (N = 27, 33%) estimated this prevalence to be <25% (**eFigure 1**). Respondents’ estimates of delirium prevalence differed by hospital (χ^2^ = 6.52, p = 0.04), where respondents from AMCs reported a higher delirium prevalence than those in community non-teaching hospitals (50% vs. 30%). Most respondents (74%) agreed that hypoactive and mixed hyperactive-hypoactive delirium are more prevalent than hyperactive delirium. Nearly all (90%) respondents felt that delays in recognition and treatment of ICU delirium prolong ICU stay and lead to worse long-term clinical outcomes.

**Table 2.**
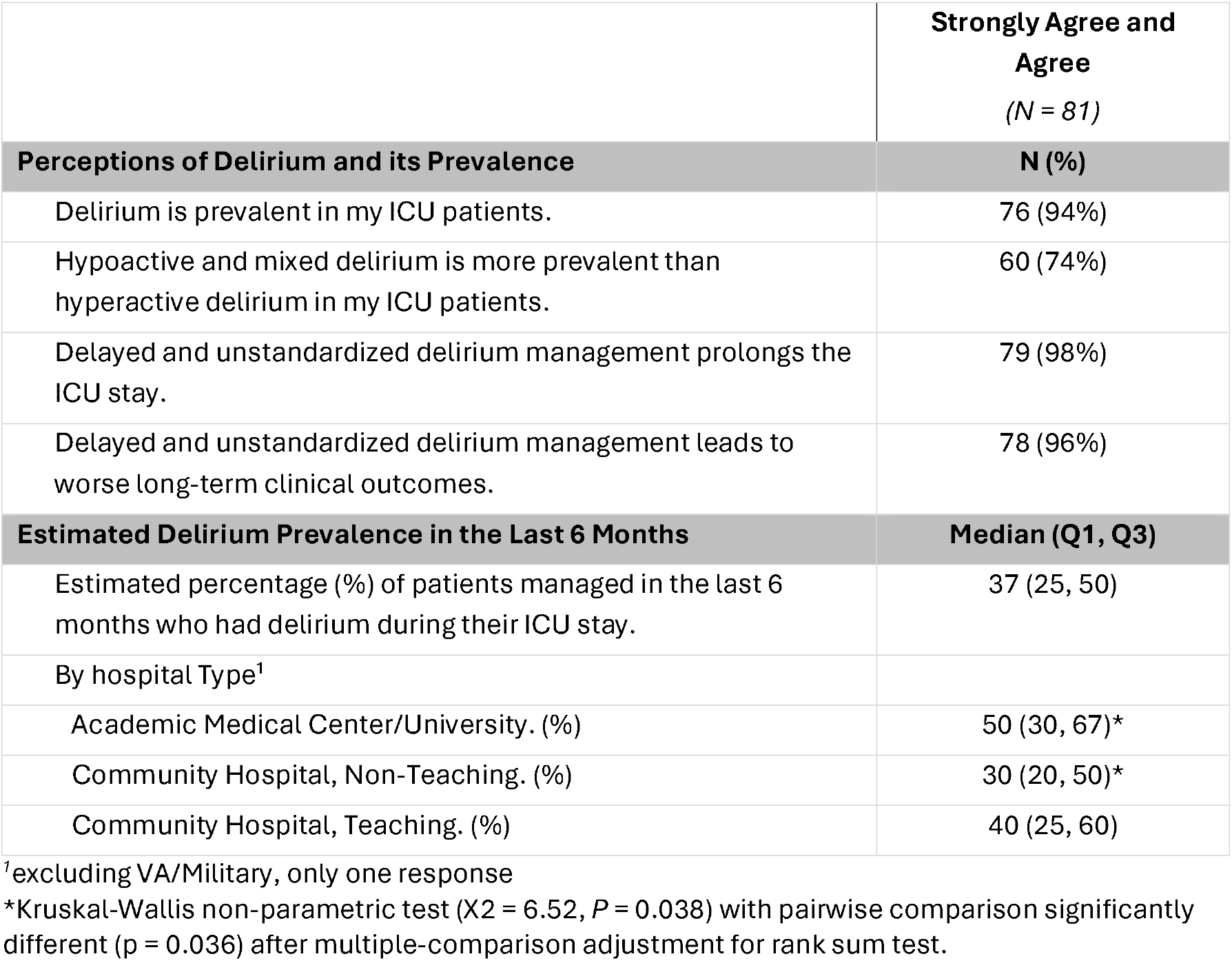
Perceptions of ICU delirium prevalence and detection.

**Table 3** summarizes respondents’ perceived delirium assessment and recognition practices, and perceived limitations of these practices. The majority of respondents (79%) reported they use the CAM-ICU performed by bedside nurses to assess patients for delirium, but 72% (N=46) of these also reported they rely on a diagnosis of delirium by the intensivist. Most (63%) reported that delirium assessments with a validated screening tool like the CAM-ICU or the ICDSC are conducted at least twice daily in their ICUs, but 40% (N = 33) perceived these assessments are completed appropriately less than 50% of the time. The majority of respondents agreed with the statement that bedside assessments poorly detect delirium in patients with hypoactive delirium (84%), in invasively mechanically ventilated patients regardless of their sedation level (90%), and in patients with acute neurologic conditions other than delirium (e.g., non-convulsive seizures, stroke) (84%). A majority (63%) agreed that current delirium assessments delay recognition of delirium, which leads to delays in effective delirium management efforts in their ICUs.

**Table 3.** Reported delirium assessment and practices, and their perceived limitations.

|  | Strongly Agree and Agree <sup>1</sup><br>(N = 81) |
| --- | --- |
| **Current Delirium Assessment Practices** |  |
| How is delirium assessed and diagnosed in your practice?<br>(Multiple selections allowed) |  |
| CAM-ICU assessment by nurses. | 64 (79%) |
| Other delirium assessment by nurses (e.g., ICDSC). | 13 (16%) |
| Diagnosis by physician, provider dependent. | 62 (77%) |
| Do not really assess delirium. | 4 (5%) |
| Others. | 1 (1%) |
| Number of Delirium Assessments per Day |  |
| >2x | 10 (12%) |
| 2x | 41 (51%) |
| 1x | 17 (21%) |
| ad-hoc | 13 (16%) |
| Percentage of Reliable Assessments |  |
| <20% | 10 (12%) |
| 20-50% | 23 (28%) |
| 51-80% | 35 (43%) |
| >80% | 13 (16%) |
| **Perceived Limitations of Current Delirium Assessment Tools (e.g., CAM-ICU, ICDSC)** |  |
| They poorly recognize delirium in non-verbal intubated patients (regardless of level of sedation). | 73 (90%) |
| They poorly recognize delirium in patients at a minimal level of sedation (RASS = -1 to 0). | 58 (72%) |
| They poorly recognize delirium in patients with light sedation (RASS -2). | 70 (86%) |
| They poorly recognize delirium in patients with moderate sedation (RASS -3). | 72 (89%) |
| They poorly recognize hypoactive delirium. | 68 (84%) |
| They poorly recognize and differentiate delirium in patients with an acute neurologic condition (e.g., acute stroke, non-convulsive seizures). | 68 (84%) |
| They delay the recognition of delirium, leading to delays in effective delirium reduction/management efforts in my ICU. | 51 (63%) |

<sup>1</sup>n (%)
CAM-ICU: Confusion Assessment Method for the Intensive Care Unit
ICDSC: Intensive Care Delirium Screening Checklist

### Delirium Management Practices

Respondents unanimously agreed that early recognition and management of ICU delirium using a standardized delirium management protocol would improve clinical outcomes, and the majority (96%) agreed that improving delirium recognition and treatment in ICU patients should be a higher priority in their ICUs (**Table 4**). Yet only 25% reported they currently use a standardized delirium management protocol or decision tree. Frequently reported non-protocolized approaches to managing ICU delirium included a provider review of sedative choice and use of deliriogenic medications, vital signs, and other physiologic parameters and lab results to rule out cardiopulmonary, metabolic or infectious causes (**eFigure 2**). Of note, there was a high degree of variability in the reported choice of sedatives used, with most respondents (58%) reporting that 20% or more of their patients receive benzodiazepines. Protocolization and sedative use were not associated with respondent profession or hospital type (p= 0.08 and p = 0.10, respectively, **eTable 1**). Respondents unanimously agreed that employing non-pharmacologic delirium management strategies can shorten ICU length of stay and improve outcomes in patients. Yet only 26% reported they currently utilize non-pharmacologic interventions (e.g., promoting sleep, mobility, and family visitation) to manage delirium, and only 19% of respondents reported routinely tracking ICU delirium quality metrics (e.g., frequency of completed delirium assessments, proportion of positive delirium assessments, delirium-free days, etc.).

**Table 4.** Perceptions on current delirium management and unmet needs in delirium recognition and management.

|  | **Responses <sup>1</sup>**<br>(N = 81) |
| --- | --- |
| **Current Delirium Management Practices** |  |
| Standard Patient Management Protocol or Decision Tree are in Place. | 20 (25%)<sup>2</sup> |
| Delirium Assessment (e.g., CAM-ICU) is Implemented in ICU Protocol or Standard ICU Orders. | 60 (74%)<sup>2</sup> |
| ICU Patients on Benzodiazepines (%), *median (Q1, Q3)*. | 20% (10%, 40%) |
| Delirium-related Metrics are Tracked in the Department. | 15 (19%)<sup>2</sup> |
| **Perceived Unmet Needs in Delirium Recognition and Management** |  |
|  | **Agree and Strongly Agree** |
| I believe improving the recognition of delirium in my ICU should be a higher priority in daily practice. | 78 (96%) |
| The use of early and standardized delirium management strategies will improve the post-ICU outcome(s) of my patients. | 81 (100%) |
| Delirium management strategies are important to use regardless of patients' level of sedation. | 78 (96%) |
| Even though there is not a single effective drug, there are things I can do to manage delirium and improve patient outcomes. | 81 (100%) |
| Delirious patients need to be managed as soon as delirium is recognized (e.g., within an hour). | 69 (85%) |
| Current delirium assessment approaches / tools (e.g., CAM-ICU) are adequate to evaluate the effectiveness of delirium management strategies in my patients. | 46 (57%) |

<sup>1</sup>n (%); Median (Q1, Q3)
<sup>2</sup>"Yes" responses
ICU: Intensive Care Unit; CAM-ICU: Confusion Assessment Method for the ICU

The majority (85%) agreed that newly-delirious patients should be managed as soon as possible (e.g., within first hour of diagnosis), and nearly all (96%) agreed that delirium management strategies are important to use regardless of patient’s sedation level and that delirium recognition should be a higher priority in their ICU (**Table 4**). However, only half of the respondents (57%) agreed that current delirium assessments and tools are adequate to evaluate the effectiveness and management of delirium strategies in their patients.

### Potential Role of Monitoring Technologies

Nearly all respondents (93% for Option 1 vs. 94% for Option 2) agreed that a reliable objective delirium monitor that provides either a binary detection or a continuous output display would be clinically valuable. Moreover, they agreed that a binary delirium detector or continuous delirium monitor could provide timely and actionable information to accelerate and improve delirium management in ICU patients; 89% and 90%, respectively (**Table 5**). Most respondents (84% for Option 1 vs. 86% for Option 2) agreed that the use of a continuous delirium monitor would also improve ICU patients’ outcomes.

**Table 5.**
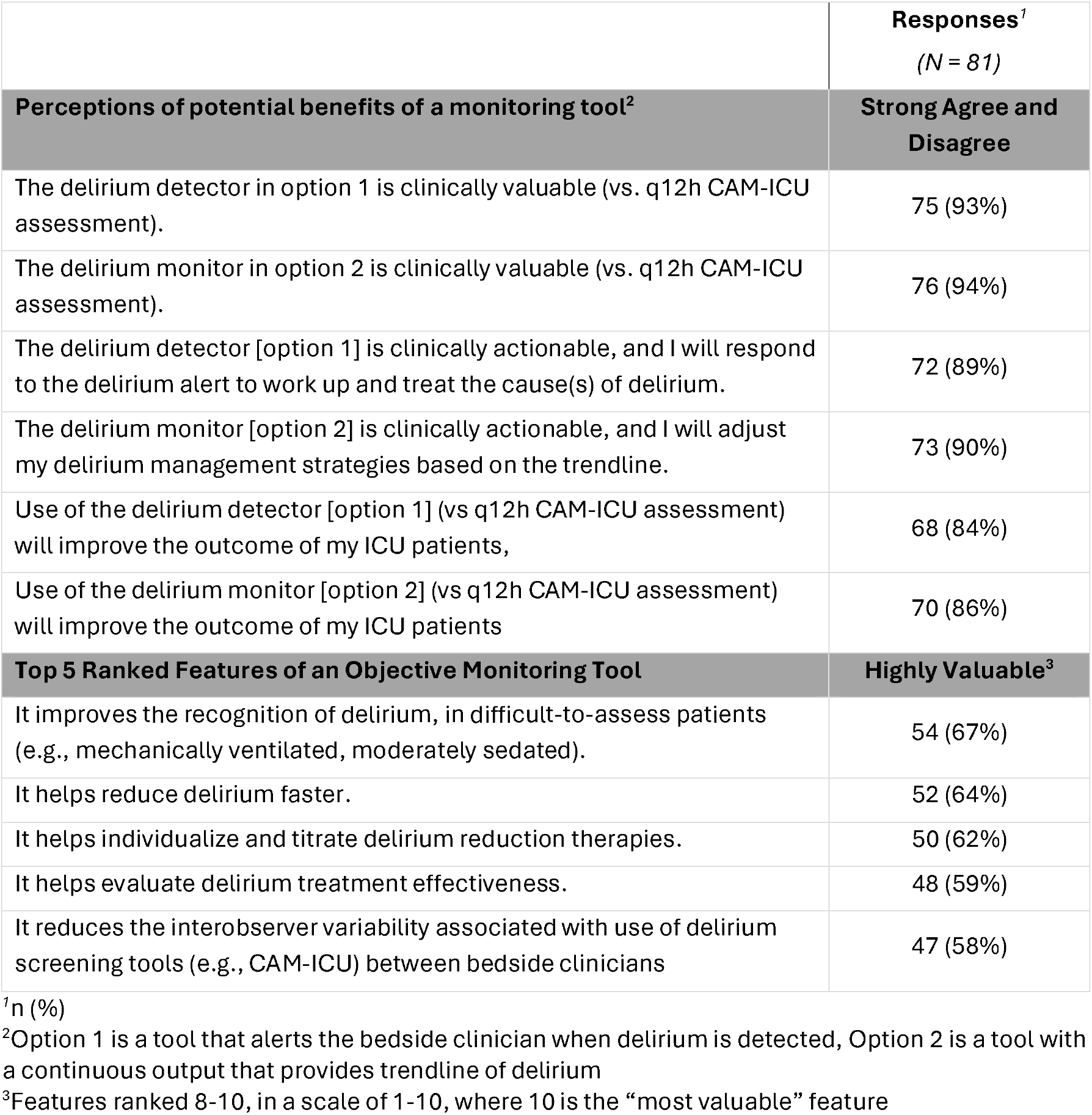
Perceptions of potential benefits of an objective monitoring tool, and valuable features.

When asked which features of a continuous delirium monitor would be most valuable (**Table 5** and **eFigure 3**), more than 60% of respondents reported the following features would be highly valuable: improving recognition of delirium in difficult-to-assess patients, helping to recognize delirium faster, and helping to individualize and titrate delirium reduction therapies. There were no significant differences in these responses either by respondent profession or hospital type. However, there was a trend for respondents from community hospitals, both teaching and non-teaching, to value tools that would help reduce the use of delirium treatment medications when compared to respondents from AMC or university hospitals (63% vs. 47% vs. 26%, p = 0.05, **eTable 2**), and for nurses to value more than physicians a tool that would reduce the bedside nursing burden associated with (CAM-ICU) delirium assessment (68% vs. 44%, p = 0.06, **eTable 2**)

## Discussion

This survey is the first of its kind, to our knowledge, to examine ICU leaders’ perceptions of the challenges associated with monitoring, assessing and managing ICU delirium. The key findings of this survey are that ICU leaders believe that: 1) there are significant clinical limitations to bedside delirium assessments for detecting ICU delirium in a timely manner, 2) these tools fall short for detecting delirium in certain patient populations, 3) current bedside tools do not effectively evaluate the impact of delirium interventions strategies, and 4) an objective delirium monitoring tool, be it a binary detector or one with a continuous output, could potentially improve delirium detection, management and outcomes for critically ill patients.

Although perceptions among leaders of delirium prevalence in their ICU mostly align with published pooled prevalences^1,5,20^, around one-third of respondents reported lower than average ICU delirium prevalence in the last six months. This could be due to the type of ICUs they manage and the age and illness severity of their patient population. However, nearly half of respondents believed the delirium assessments currently used in their ICUs are reliably completed less than half the time. Moreover, a majority of respondents perceived that current tools delay recognition of delirium. Taken together, this suggests a perceived under-recognition of delirium in the ICUs surveyed, potentially due to either limitations of current assessment tools or inadequate education and implementation practices.

We also found that respondents lack confidence in current delirium assessments, like the CAM-ICU and ICDSC, because they perceive these tools are inadequate for detecting hypoactive delirium or delirium in non-verbal or sedated patients. These perceptions align well with previously reported delirium detection barriers by nursing personnel, where even after education, confidence in performing delirium assessments in non-verbal intubated patients remained low^21,22^. Our results indicate that ICU leaders also recognize these barriers and perceived similar challenges of implementing an assessment in real practice and sustaining the quality of the assessments^23^.

Overall, there was high agreement about the perceived limited effectiveness of current assessments to evaluate whether delirium management strategies are working. This could be related to current assessments only representing a snapshot in time about patient status, while ICU delirium often fluctuates over the course of hours and days^1,24^. Our findings of reported low trust in the assessments mirrors reports from some non-US studies^25^, and it may have implications about the type of strategies implemented, despite the existence of validated strategies to manage and prevent delirium^26,27^.

When considering theoretical objective delirium monitors, respondents had very high agreement that such tools would be clinically valuable, that the information could be clinically actionable, and that it could improve patients’ outcomes. Although both of the options described in our survey (i.e., a binary delirium detector and another with a continuous output) were considered as potentially valuable by the majority of respondents, the respondents slightly favored monitoring technologies able to provide a continuous output and with the potential to help individualize delirium treatment interventions. Moreover, most respondents reported that the most valuable features for an objective delirium monitor would be its ability to improve the recognition of delirium in mechanically ventilated or other difficult-to-assess patients. While the idea of an objective monitoring tool may appear favorable to ICU leaders, opinions of bedside providers may differ, especially since one of the features that was rated lower priority was the ability of the tool to reduce burden on nursing staff. This may be at odds with persistent nursing-related challenges following the COVID-19 pandemic^28^. To this end, future technological interventions or monitoring tools should be evaluated by physicians and nurses alike, ideally in collaboration. Given early evidence that technology-aided interventions and digital-based tools may help nursing adherence to guidelines^29^, it is possible that technologies with more clinically interpretable output could be successfully incorporated in ICU workflows without increasing nursing burden and potentially impacting delirium care. Further, the type of devices surveyed could potentially help raise awareness of how prevalent ICU delirium actually is, and to take corrective actions to help reduce their delirium burden.

Our study has several limitations. Perceptions of ICU leaders may not align with the actual delirium assessment and treatment practices, workflows, or protocols used in each respondent’s ICU. Our respondent sample was unbalanced, with around two-thirds identifying as physicians, so answers may not generalize well to other ICU personnel. The nurse and physician leaders surveyed were not from the same hospital, so any differences between the two professions could be attributed to the suspected variability in delirium practices between hospitals. The results we report may not represent the perceptions of ICU leaders not surveyed, and the survey deployment method we used did not allow us to track a response rate. Although we explored the impact of hospital type as well as profession, our analyses may be underpowered and other characteristics of the hospitals not captured in the survey could impact the perceptions we report. The assessment tools given as an example in our survey were the CAM-ICU and ICDSC, which may limit the generalizability of our results. However, in a sub-analysis of the World Delirium Awareness Day survey of 91 units across the US, the majority of ICUs surveyed reported using CAM-ICU as their standard assessment tool^13^. Finally, our survey was conducted with an industry sponsor and was initially designed for market research, although the participants were blind to any company or brand name and the questions were developed in partnership between researchers and sponsors.

## Conclusions

Physician and nurse ICU leaders across AMC and community hospitals in the US lack confidence in current delirium assessments, particularly when used to assess intubated, sedated, and non-verbal ICU patients. They agree that an objective continuous delirium monitor may provide actionable and clinically valuable information that could address these shortcomings. Our results highlight important areas for developing and researching delirium monitoring technologies.

## Data Availability

All data produced in the present study are not publicly available and are the property of Ceribell.

## Acknowledgements

We thank John Rickelman Jr., DO. for his feedback on the manuscript.

## References

1. Wilson JE, Mart MF, Cunningham C, et al. Delirium. Nature Reviews Disease Primers. 2020/11/12/ 2020;6(1):90. doi:10.1038/s41572-020-00223-4

2. Shehabi Y, Riker RR, Bokesch PM, Wisemandle W, Shintani A, Ely EW. Delirium duration and mortality in lightly sedated, mechanically ventilated intensive care patients. Crit Care Med. Dec 2010;38(12):2311–8. doi:10.1097/CCM.0b013e3181f85759

3. Pisani MA, Kong SY, Kasl SV, Murphy TE, Araujo KL, Van Ness PH. Days of delirium are associated with 1-year mortality in an older intensive care unit population. Am J Respir Crit Care Med. Dec 1 2009;180(11):1092–7. doi:10.1164/rccm.200904-0537OC

4. Pandharipande PP, Girard TD, Jackson JC, et al. Long-term cognitive impairment after critical illness. N Engl J Med. Oct 3 2013;369(14):1306–16. doi:10.1056/NEJMoa1301372

5. Leong AY, Edginton S, Lee LA, et al. Prevalence and incidence of ICU delirium and pain: a systematic review and meta-analysis. Intensive Care Medicine. 2025/11/01/ 2025;51(11):2093–2103. doi:10.1007/s00134-025-08167-7

6. Marra A, Ely EW, Pandharipande PP, Patel MB. The ABCDEF Bundle in Critical Care. Critical Care Clinics. 2017;33(2):225–243. doi:10.1016/j.ccc.2016.12.005

7. Ely EW, Inouye SK, Bernard GR, et al. Delirium in Mechanically Ventilated PatientsValidity and Reliability of the Confusion Assessment Method for the Intensive Care Unit (CAM-ICU). JAMA. 2001;286(21):2703–2710. doi:10.1001/jama.286.21.2703

8. Ely EW, Margolin R, Francis J, et al. Evaluation of delirium in critically ill patients: validation of the Confusion Assessment Method for the Intensive Care Unit (CAM-ICU). Crit Care Med. Jul 2001;29(7):1370–9. doi:10.1097/00003246-200107000-00012

9. Bergeron N, Dubois MJ, Dumont M, Dial S, Skrobik Y. Intensive Care Delirium Screening Checklist: evaluation of a new screening tool. Intensive Care Med. May 2001;27(5):859–64. doi:10.1007/s001340100909

10. Ista E, Redivo J, Kananur P, et al. ABCDEF Bundle Practices for Critically Ill Children: An International Survey of 161 PICUs in 18 Countries*. Critical Care Medicine. 2022;50(1):114–125. doi:10.1097/ccm.0000000000005168

11. Moraes FdS, Marengo LL, Moura MDG, et al. ABCDE and ABCDEF care bundles: A systematic review of the implementation process in intensive care units. Medicine. 2022;101(25):e29499. doi:10.1097/md.0000000000029499

12. Thomas AG, Davis SP. Facilitators and Barriers for Nurses Implementing Delirium PADIS Guidelines in the Intensive Care Unit: An Integrative Review. Dimens Crit Care Nurs. Nov-Dec 01 2025;44(6):328–352. doi:10.1097/dcc.0000000000000725

13. Lindroth H, Byrnes T, Fuchita M, et al. Delirium in the United States: Results From the 2023 Cross-Sectional World Delirium Awareness Day Prevalence Study. J Acad Consult Liaison Psychiatry. Sep-Oct 2024;65(5):417–430. doi:10.1016/j.jaclp.2024.06.005

14. Lindroth H, Liu K, Szalacha L, et al. World delirium awareness and quality survey in 2023-a worldwide point prevalence study. Age Ageing. Nov 1 2024;53(11) doi:10.1093/ageing/afae248

15. Ragheb J, Norcott A, Benn L, et al. Barriers to delirium screening and management during hospital admission: a qualitative analysis of inpatient nursing perspectives. BMC Health Services Research. 2023/06/29/ 2023;23(1):712. doi:10.1186/s12913-023-09681-416.

16. Nydahl P, Liu K, Bellelli G, et al. A world-wide study on delirium assessments and presence of protocols. Age and Ageing. 2024;53(7) doi:10.1093/ageing/afae129

17. van der Kooi AW, Rots ML, Huiskamp G, et al. Delirium Detection Based on Monitoring of Blinks and Eye Movements. The American Journal of Geriatric Psychiatry. 2014/12/01/ 2014;22(12):1575–1582. doi:10.1016/j.jagp.2014.01.001

18. Van Der Kooi AW, Zaal IJ, Klijn FA, et al. Delirium detection using EEG. Chest. 2015 2015;147(1):94–101. doi:10.1097/WNP.0000000000000049

19. Tang E, Laverty M, Weir A, et al. Development and feasibility of a smartphone-based test for the objective detection and monitoring of attention impairments in delirium in the ICU. Journal of Critical Care. 2018/12/01/ 2018;48:104–111. doi:10.1016/j.jcrc.2018.08.01920.

20. Krewulak KD, Stelfox HT, Leigh JP, Ely EW, Fiest KM. Incidence and Prevalence of Delirium Subtypes in an Adult ICU: A Systematic Review and Meta-Analysis*. Critical Care Medicine. 2018;46(12):2029–2035. doi:10.1097/ccm.0000000000003402

21. Mailhot T, Crump L, Clausen C, et al. Nurse-perceived barriers and facilitators influencing optimal delirium care in acute care patients: Findings from the INVOLVE_RN study on barriers and facilitators. Geriatric Nursing. 2025/05/01/ 2025;63:76–84. doi:10.1016/j.gerinurse.2025.03.023

22. Law TJ, Leistikow NA, Hoofring L, Krumm SK, Neufeld KJ, Needham DM. A survey of nurses’ perceptions of the intensive care delirium screening checklist. Dynamics. 2012 2012;23(4):18–24.

23. Numan T, van den Boogaard M, Kamper AM, et al. Recognition of Delirium in Postoperative Elderly Patients: A Multicenter Study. J Am Geriatr Soc. 2017/09/01 2017;65(9):1932–1938. doi:10.1111/jgs.14933

24. The DSM-5 criteria, level of arousal and delirium diagnosis: inclusiveness is safer. BMC Med. Oct 8 2014;12:141. doi:10.1186/s12916-014-0141-2

25. Oxenbøll-Collet M, Egerod I, Christensen V, Jensen J, Thomsen T. Nurses’ and physicians’ perceptions of Confusion Assessment Method for the intensive care unit for delirium detection: focus group study. Nursing in Critical Care. 2018/01/01 2018;23(1):16–22. doi:10.1111/nicc.12254

26. Pun BT, Balas MC, Barnes-Daly MA, et al. Caring for Critically Ill Patients with the ABCDEF Bundle: Results of the ICU Liberation Collaborative in Over 15,000 Adults. Crit Care Med. Jan 2019;47(1):3–14. doi:10.1097/ccm.0000000000003482

27. Barr J, Paulson SS, Kamdar B, et al. The Coming of Age of Implementation Science and Research in Critical Care Medicine. Crit Care Med. Aug 1 2021;49(8):1254–1275. doi:10.1097/ccm.0000000000005131

28. Potter KM, Pun BT, Maya K, et al. Delirium and Coronavirus Disease 2019: Looking Back, Moving Forward. Crit Care Nurs Clin North Am. Sep 2024;36(3):415–426. doi:10.1016/j.cnc.2023.12.003

29. Al Siyabi A, Peddle M, Tomlinson EJ. Exploring the use of digital technologies in the management of delirium in acute hospital settings: A scoping review. Contemporary Nurse. 2025/10/18 2025:1–20. doi:10.1080/10376178.2025.2572342

