## Supplementary Material for "Current Gaps in Delirium Recognition and Management: A Cross-Sectional Survey of ICU Physician and Nurse Leaders"

### Supplementary Tables and Figures

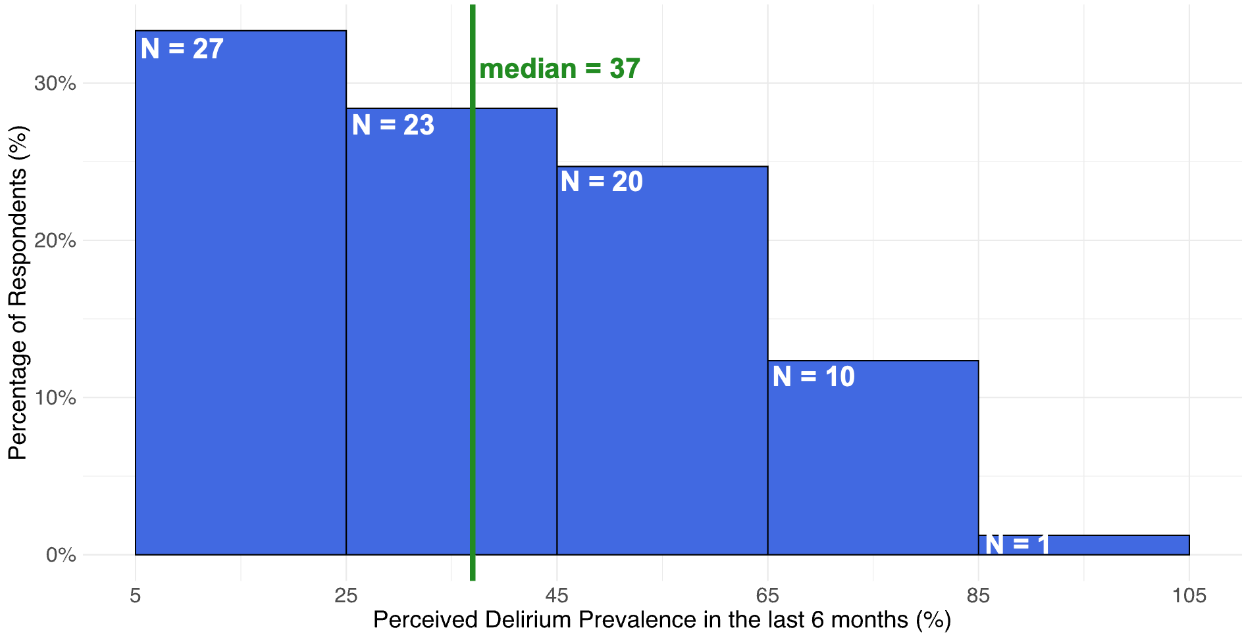


**eFigure1. Distribution of responses of the perceived delirium prevalence in the last six month across the managed ICUs**

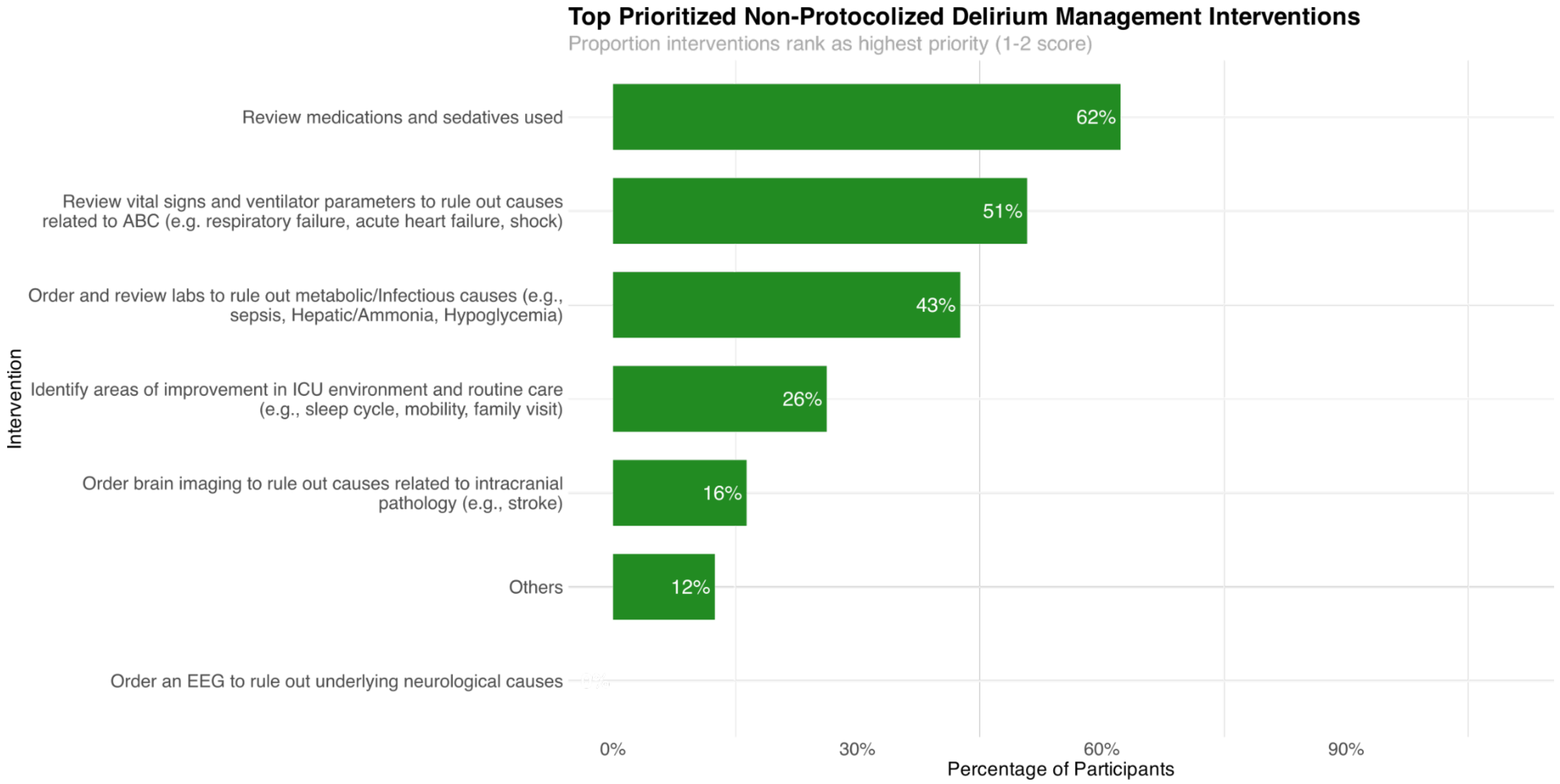


**eFigure 2. Summary of interventions ranked as top highest priorities by participants who reported not having protocolized delirium management**

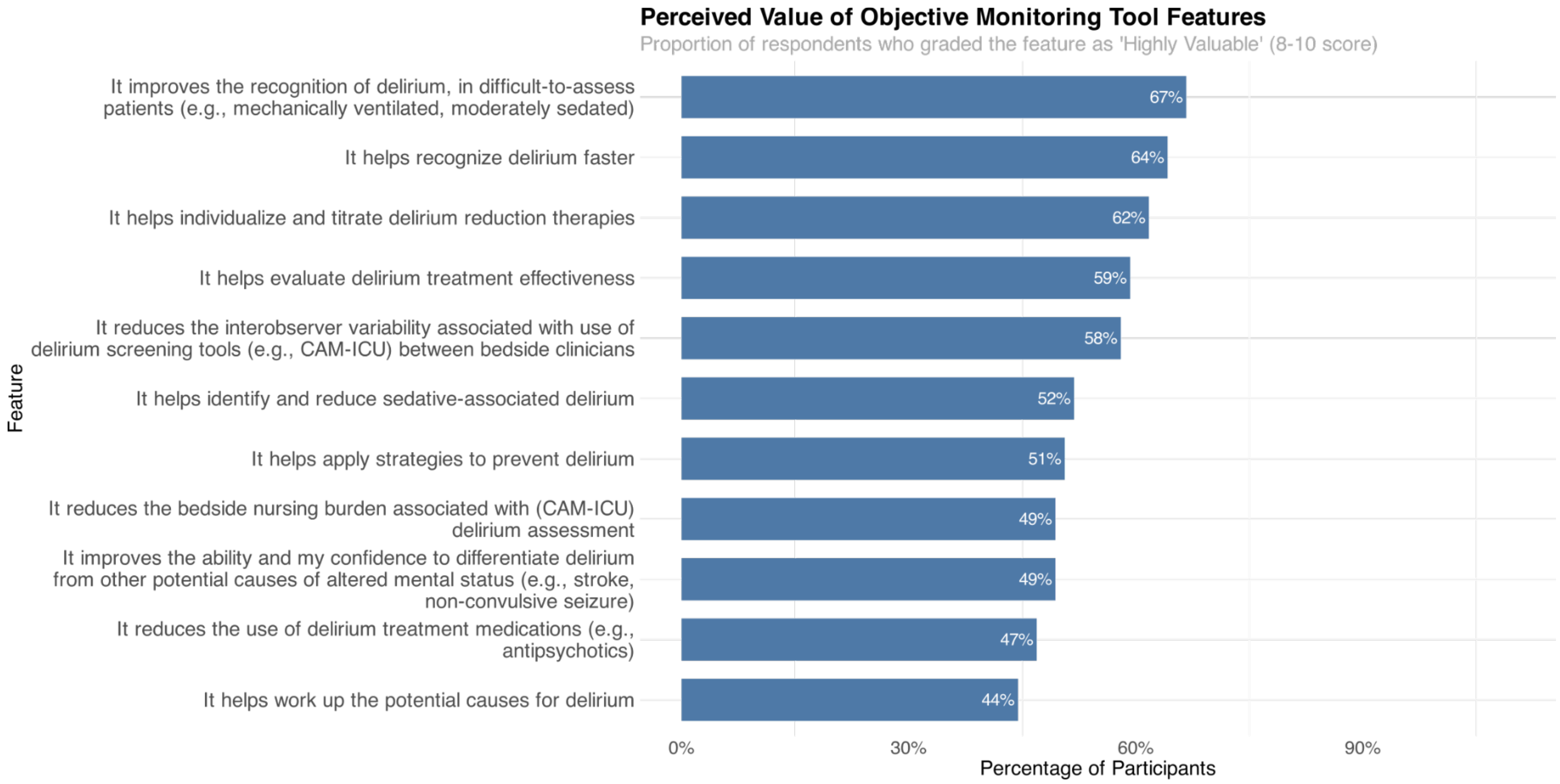


**eFigure3. Summary of all surveyed features, ranked as “most valuable” in an objective monitoring tool technology.**

**eTable 1. Perceptions on current delirium detection and management practices, with breakdown by reported hospital type and profession.**

| | Hospital type[1] | | | | Profession | | |
|---|---|---|---|---|---|---|---|
| **Features** | **AMC/University (N = 19[2])** | **Community Hospital, Non-Teaching (N = 34[2])** | **Community Hospital, Teaching (N = 27[2])** | ***P* value[3]** | **Nurse (RN/NP/Other) (N = 19[2])** | **Physician (N = 62[2])** | ***P* value[4]** |
| **Standard Patient Management Protocol or Decision Tree are in Place.** | 5 (26%) | 11 (32%) | 4 (15%) | 0.3 | 3 (16%) | 17 (27%) | 0.4 |
| **Delirium Assessment (e.g., CAM-ICU) is Implemented in ICU Protocol or Standard ICU Orders.** | 15 (79%) | 26 (76%) | 18 (67%) | 0.7 | 14 (74%) | 46 (74%) | >0.9 |
| **ICU Patients on Benzodiazepines (%).** | 10 (5, 40) | 20 (10, 35) | 30 (15, 50) | 0.10 | 30 (10, 70) | 20 (10, 40) | 0.084 |
| **Delirium-related Metrics are Tracked in the Department.** | 3 (16%) | 9 (26%) | 3 (11%) | 0.3 | 1 (5.3%) | 14 (23%) | 0.2 |

[1]Excluding single answer with VA/Military response
[2]n (%); Median (Q1, Q3)
[3]Kruskal-Wallis rank sum test
[4]Fisher's exact test

**eTable 2. Features in an objective monitoring tool that would be most valuable, broken down by hospital type and by profession.**

| | Hospital Type[1] | | | | Profession | | |
|---|---|---|---|---|---|---|---|
| **Characteristic** | **AMC/University (N = 19[2])** | **Community Hospital, Non-Teaching (N = 34[2])** | **Community Hospital, Teaching (N = 27[2])** | ***P* value[3]** | **Nurse (RN/NP/Other) (N = 19[2])** | **Physician (N = 62[2])** | ***P* value[3]** |
| **It helps recognize delirium faster** | 12 (63%) | 19 (56%) | 20 (74%) | 0.3 | 14 (74%) | 38 (61%) | 0.3 |
| **It improves the recognition of delirium, in difficult-to-assess patients (e.g., mechanically ventilated, moderately sedated)** | 12 (63%) | 21 (62%) | 20 (74%) | 0.6 | 12 (63%) | 42 (68%) | 0.7 |
| **It reduces the interobserver variability associated with use of delirium screening tools (e.g., CAM-ICU) between bedside clinicians** | 11 (58%) | 18 (53%) | 17 (63%) | 0.7 | 12 (63%) | 35 (56%) | 0.6 |

| | | | | | | | |
|---|---|---|---|---|---|---|---|
| **It improves the ability and my confidence to differentiate delirium from other potential causes of altered mental status (e.g., stroke, non-convulsive seizure)** | 10 (53%) | 16 (47%) | 13 (48%) | >0.9 | 12 (63%) | 28 (45%) | 0.2 |
| **It reduces the bedside nursing burden associated with (CAM-ICU) delirium assessment** | 9 (47%) | 18 (53%) | 12 (44%) | 0.8 | 13 (68%) | 27 (44%) | 0.058 |
| **It helps work up the potential causes for delirium** | 6 (32%) | 14 (41%) | 15 (56%) | 0.3 | 10 (53%) | 26 (42%) | 0.4 |
| **It helps evaluate delirium treatment effectiveness** | 8 (42%) | 19 (56%) | 20 (74%) | 0.086 | 13 (68%) | 35 (56%) | 0.4 |
| **It helps individualize and titrate delirium reduction therapies** | 12 (63%) | 18 (53%) | 19 (70%) | 0.4 | 13 (68%) | 37 (60%) | 0.5 |
| **It helps identify and reduce sedative-associated delirium** | 9 (47%) | 16 (47%) | 17 (63%) | 0.4 | 10 (53%) | 32 (52%) | >0.9 |

| | | | | | | | |
|---|---|---|---|---|---|---|---|
| **It reduces the use of delirium treatment medications (e.g., antipsychotics)** | 5 (26%) | 16 (47%) | 17 (63%) | 0.051 | 10 (53%) | 28 (45%) | 0.6 |
| **It helps apply strategies to prevent delirium** | 8 (42%) | 16 (47%) | 16 (59%) | 0.5 | 11 (58%) | 30 (48%) | 0.5 |

[1]Excluding single answer with VA/Military response

[2]n (%)

[3]Fisher's exact test
